# Aetiological Factors and Clinical Profile of Patients with Extreme Leukocytosis: A Retrospective Hospital-Based Study in the Southern Highland, Tanzania

**DOI:** 10.1101/2025.03.18.25324160

**Authors:** Adamu Kilungu, Anitha Mrosso

**Affiliations:** Clinical Research, Training and Consultancy Unit, Mbeya Zonal Referral Hospital; Internal Medicine Department, Mbeya Zonal Referral Hospital; Mbeya College of Health and Allied Sciences, University of Dar Es Salaam, Mbeya, Tanzania

**Author notes:** **Corresponding Author** Adamu Kilungu, Clinical Research, Training and Consultancy Unit, Mbeya Zonal Referral Hospital, P.O. BOX 419, Mbeya, Tanzania., **Email:**.

**Keywords:** Extreme leukocytosis, haematological malignancies, leukemoid reaction, severe leukocytosis, hyperleukocytosis

## Abstract

**Background:** Extreme leukocytosis (EL), defined as an abnormally high white blood cell (WBC) count, is a critical clinical indicator associated with various underlying conditions, such as infections and malignancies. This study investigated the etiological factors and clinical profiles of patients presenting with EL at Mbeya Zonal Referral Hospital (MZRH).

**Methods:** A retrospective cohort study was conducted among patients with WBC counts ≥50×10^9^/L who attended MZRH between January 2021 and December 2022. Data were retrieved from electronic health records and analyzed using Stata Version 16.

**Results:** A total of 178 patients with EL were included in the study. Malignant conditions accounted for 47.2% of cases, with haematological malignancies comprising 89.3%, predominantly chronic myeloid leukaemia (CML). Infections were the second most frequent cause (43.2%). Patients with malignancies had significantly higher median WBC counts (221 vs. 56 × 10^9^/L, p<0.0001) and were more likely to present with symptoms such as bleeding, bone pain, B symptoms, splenomegaly, hepatomegaly, and lymphadenopathy. Severe thrombocytopenia (platelet count <50 × 10^9^/L) was more common in the malignant group (p=0.0008).

**Conclusion:** Malignant etiologies, particularly haematological malignancies, are a leading cause of EL in patients with WBC counts ≥50 × 10^9^/L. Clinicians should maintain a high suspicion of malignancies in such patients and conduct thorough diagnostic evaluations to ensure optimal management.

## Introduction

Leukocytosis, defined as an elevated white blood cell (WBC) count above the normal threshold of >11×10^9^/L, is a common clinical finding. Extreme leukocytosis (EL) is characterized by a markedly elevated WBC count, often exceeding twice the upper normal limit. However, the specific cut-off to define EL remains variable across studies, ranging from >25×10^9^/L to >50×10^9^/L (Reding et al. 1998; Granger and Kontoyiannis 2009; Zachary P. Hugo, MD, Michael C. Perry, MD, David P. Steensma, MD 2008; Viner, Berger, and Bengualid 2023). EL can result from a variety of causes, including infections, haematological malignancies, corticosteroid use, growth factors, haemorrhage, and solid tumours (Reding et al. 1998; Granger and Kontoyiannis 2009; Zachary P. Hugo, MD, Michael C. Perry, MD, David P. Steensma, MD 2008).

Infections, particularly urinary tract infections and bacterial pneumonia are frequently cited as common causes of EL when lower WBC cut-offs are applied (Reding et al. 1998; Widick and Winer 2016). Clostridium difficile has also been implicated in EL, particularly in septic patients, where prolonged EL can indicate poor prognosis and increased mortality (Naaraayan et al. 2015; Lioni et al. 2018; Potasman & Grupper 2013; Asadollahi et al. 2011). Conversely, haematological malignancies, such as chronic myeloid leukaemia (CML) and acute myeloid leukaemia (AML), are strongly associated with extreme leukocytosis (Bewersdorf and Zeidan 2020; Kittivisuit et al. 2023; Christoph 2015; Gong et al. 2014). Studies suggest that higher WBC counts, particularly that> 50×10^9^/L, are more indicative of malignant rather than infectious causes (Reding et al. 1998; Lawrence et al. 2007).

The administration of corticosteroids and granulocyte colony-stimulating factor (G-CSF) are also known to induce EL by causing demargination of neutrophils or promoting granulocyte production (Widick & Winer 2016; Karakonstantis et al. 2019). In the context of solid malignancies, paraneoplastic leukemoid reactions have been reported, often driven by tumour secretion of growth factors (Cvitkovic et al. 1993; Mukhopadhyay et al. 2004; Ferrer et al. 1999; Granger and Kontoyiannis 2009; McCoach et al. 2015). EL in solid malignancies is frequently associated with poor prognosis (Kasuga et al. 2001; Izard et al. 2015).

Despite its clinical importance, data on the etiologies of EL in Africa, including Tanzania, are scarce. This study addresses this gap by analyzing the etiological factors and clinical profiles of patients with EL presenting to a tertiary hospital in Tanzania.

## Methods

### Study Setting

The study was conducted at Mbeya Zonal Referral Hospital (MZRH), a tertiary healthcare facility and teaching hospital affiliated with the University of Dar es Salaam in southern Tanzania. MZRH serves approximately 10.3 million people across seven regions: Mbeya, Njombe, Katavi, Rukwa, Ruvuma, Songwe, and Iringa. The hospital has a bed capacity of 826.

### Study Design and Population

This retrospective cohort study included all patients with WBC counts ≥50×10^9^/L who presented to MZRH between January 2021 and December 2022. Patients with a prior history of haematological malignancies were excluded.

### Data Collection

Data were extracted from the hospital’s electronic medical records, including demographic information, clinical presentations, underlying conditions, and etiologies of EL. A structured case report form was used for data abstraction.

### Diagnostic Criteria

Clinical signs such as fever, positive cultures, or symptom resolution with antibiotic therapy were considered for infectious etiologies. Malignancies were diagnosed using flow cytometry and, where available, molecular testing for BCR-ABL/Philadelphia chromosome in CML cases. In the absence of advanced diagnostics, peripheral smear and bone marrow biopsy results interpreted by a consultant haematologist were used.

### Statistical Analysis

Data were analyzed using Stata Version 16. Categorical variables were expressed as frequencies and percentages, while continuous variables were summarized as means (standard deviation) or medians (interquartile range). Comparisons between groups were conducted using Chi-square or Fisher’s exact tests for categorical variables and t-tests or Wilcoxon rank-sum tests for continuous variables. A p-value of <0.05 was considered statistically significant. Multivariate logistic regression was performed to identify independent predictors of haematological malignancies.

### Ethical Considerations

Ethical approval was obtained from the Mbeya Medical Research and Ethics Committee (Ref No: SZEC-2439/R.A/24/01). All patient data were anonymized to protect confidentiality.

## Results

### Patient Demographics and Clinical Characteristics

A total of 178 patients with extreme leukocytosis (EL), defined as a WBC count of ≥50×10^9^/L, were included in the study. The median age of the patients was 19.5 years (range: 0-79 years), with 53% being male and 47% female. A significant proportion (30.3%) of patients had comorbid conditions, with sickle cell disease (SCD) being the most prevalent (77.8% of patients with comorbidities).

**Table 1.** Patient Characteristics.

**Patient Characteristics (Table 1)**
|  | **NO.** | **PERCENT (%)** |
| --- | --- | --- |
| Age-Median (Y), Range | 19.5 (0 – 79) |  |
| Males | 95 | 53% |
| **Comorbidities** | 54 | 30.3% |
| • Sickle cell disease | 42 | 77.8% |
| • PUD | 2 | 3.7% |
| • Liver Cirrhosis | 2 | 3.7% |
| • HIV/AIDS | 1 | 1.9% |
| • Tuberculosis | 2 | 3.7% |
| • CKD | 3 | 5.6% |
| • HHD | 2 | 3.7% |
| **Solid Malignancies** | 11 | 6.2% |
| • HCC | 2 | 18.2% |
| • Breast Ca | 1 | 9.1% |
| • Cervica Ca | 1 | 9.1% |
| • Chocholangiocarcinoma | 1 | 9.1% |
| • Bladder Ca | 2 | 18.2% |
| • Chondrosarcoma | 1 | 9.1% |
| • Retinoblastoma | 1 | 9.1% |
| • Willm's Tumour | 2 | 18.2% |

### Etiological Factors

Among the 178 patients, malignant conditions accounted for 47.2% of cases, with 89.3% being haematological malignancies, mainly chronic myeloid leukaemia (CML). Infections were the second most common cause of EL, observed in 43.2% of cases. Other causes, including steroid use and haemorrhage, were responsible for 9.6% of cases.

Patients with malignant conditions had significantly higher median WBC counts compared to those with infections (221×10^9^/L vs. 56×10^9^/L, p<0.0001). Furthermore, severe thrombocytopenia (platelet count <50×10^9^/L) was more frequently observed in the malignant group compared to the infection group (p=0.0008).

### Haematological Parameters

The haematological profile of patients varied between groups. The median haemoglobin (Hb) level was significantly lower in patients with infections (6.6 g/dL) compared to those with malignancies (8.0 g/dL, p=0.03). Additionally, thrombocytopenia (platelet count <150×10^9^/L) was more prevalent in the malignant group, with severe thrombocytopenia (platelet count <50×10^9^/L) being particularly common in patients with haematological malign.

**Figure 1.**
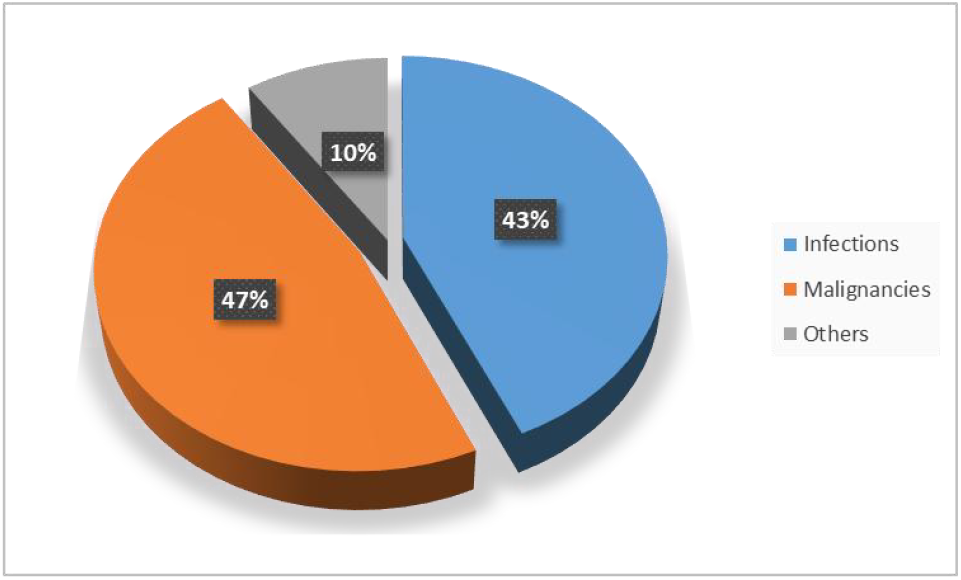
Etiologies of Extreme Leukocytosis.

**Table 2.** Haematological Parameters.

Haematological Parameters (Table 2)
| Parameter | Haematological Malignancy | Infection | P-Value |
| --- | --- | --- | --- |
| WBCs Median (IQR) $10^9/L$ | 221 (50.5 – 812) | 56 (50 – 340) | <0.0001 |
| Haemoglobin (IQR), g/dl | 8.0 (2.3 – 17.2) | 6.6 (1.5 – 17.5) | 0.03 |
| Platelets (IQR), $10^9/L$ | 151 (5 – 1771) | 276 (1 – 905) | 0.0066 |

**Figure 2.**
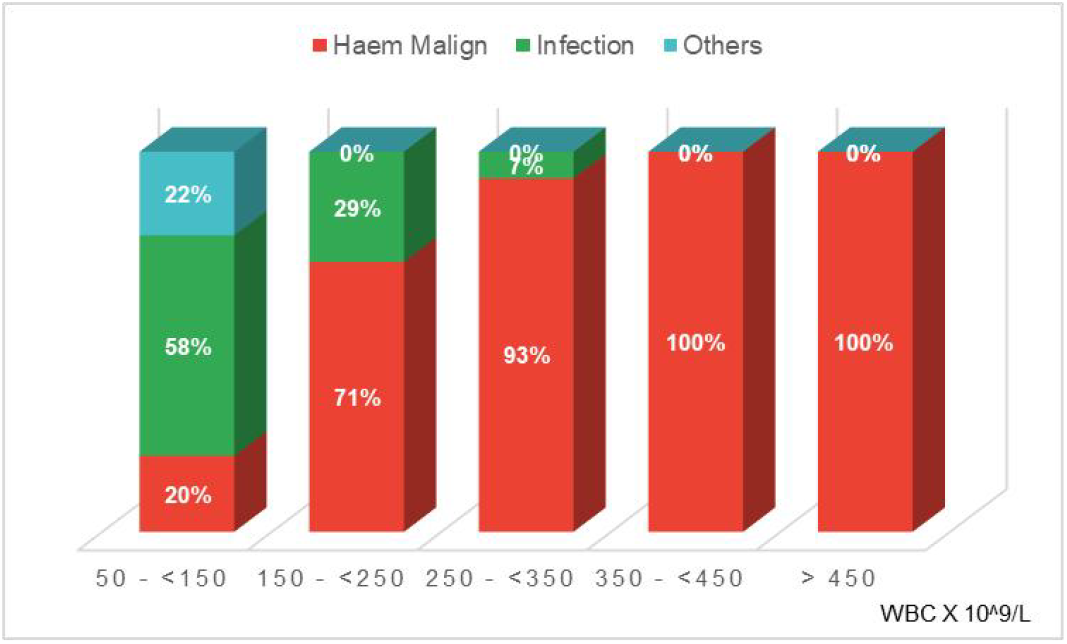
Correlation Between WBC Count and Etiology. illustrates that higher WBC counts were more likely to be associated with malignant etiologies.

**Figure 3.**
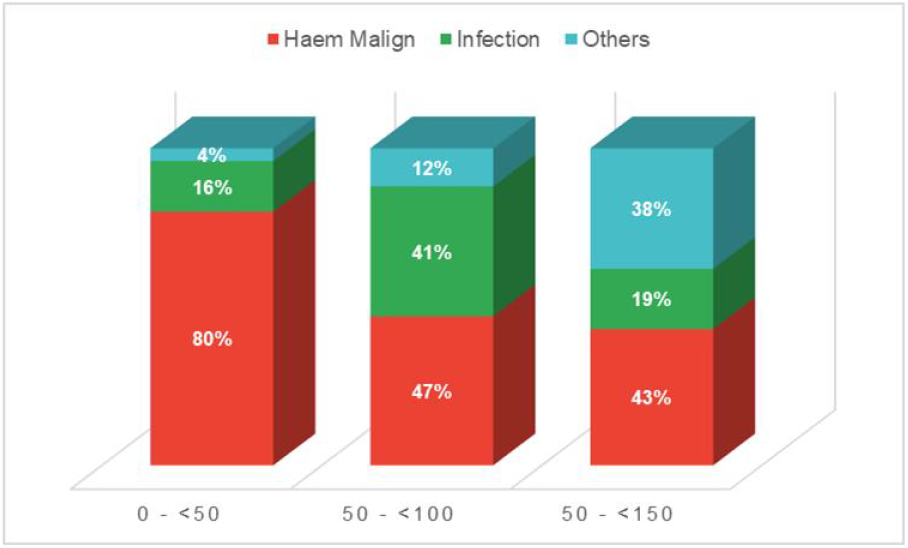
Association of Severe Thrombocytopenia with Hematological Malignancy. highlights a significant association between severe thrombocytopenia (platelet count < 50 x 10^9/L) and haematological malignancies (p=0.0008).

### Clinical Presentations

Patients with malignant conditions presented more frequently with systemic symptoms such as bleeding, bone pain, B symptoms (fever, night sweats, weight loss), splenomegaly, hepatomegaly, and lymphadenopathy. Conversely, fever was more common among patients with infections.

**Figure 4.**
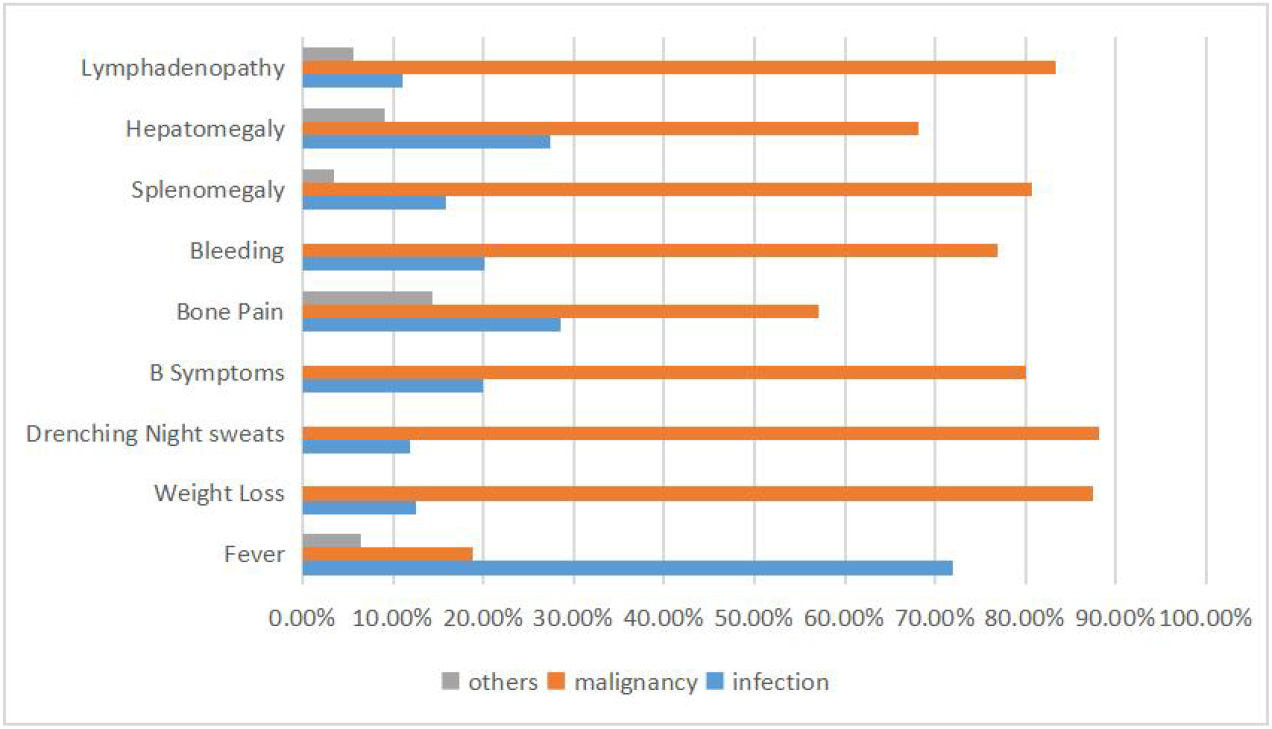
Patient Clinical Profiles.

## Discussion

This study provides important insights into the etiological factors and clinical profiles of patients with extreme leukocytosis (EL) at a tertiary hospital in Southern Tanzania. Our findings demonstrate that EL is more frequently associated with malignant etiologies, particularly haematological malignancies, compared to infections when a WBC count cut-off of ≥50×10^9^/L is used.

Haematological malignancies, mainly chronic myeloid leukaemia (CML), accounted for most EL cases (89.3%). This is consistent with previous studies that have reported a strong association between high WBC counts and Chronic myeloid leukaemia, in particular, is well known for its association with marked leukocytosis. Our study found that patients with malignancies had significantly higher median WBC counts compared to those with infections (221×10^9^/L vs. 56×10^9^/L), further supporting the idea that extreme elevations in WBC counts are more likely to be indicative of malignant processes(Widick and Winer 2016).

While infections were the second most common cause of EL in our study (43.2% of cases), they were more likely to be associated with lower WBC counts. This finding aligns with other studies using lower WBC cut-offs (e.g., 25×10^9^/L to 30×10^9^/L) and found infections to be the predominant cause (Lawrence et al. 2007). The relatively lower WBC counts in patients with infections suggest that clinicians should consider infections when the WBC count is elevated but not extreme. In contrast, a high index of suspicion for malignancy is warranted when WBC counts exceed 50×10^9^/L.

The clinical profiles of patients with EL differed based on the underlying aetiology. Patients with malignant conditions were more likely to present with symptoms such as bleeding, bone pain, and B symptoms (fever, night sweats, weight loss), as well as splenomegaly, hepatomegaly, and lymphadenopathy. These findings are consistent with prior reports highlighting these symptoms as characteristic of haematological malignancies (American Academy of Family Physicians 2024; Merck Manual 2024; MSD Manual 2024). In contrast, fever was more commonly observed in patients with infections, a finding that aligns with infection-related leukocytosis.

Severe thrombocytopenia (platelet count <50×10^9^/L) was significantly more common in patients with malignant conditions (p=0.0008). This finding suggests that thrombocytopenia, particularly when severe, may be a valuable marker for distinguishing between malignant and non-malignant causes of EL. Prior studies have also identified thrombocytopenia as a significant feature of haematological malignancies (Widick and Winer 2016; Chalasani et al. 1998).

### Implications for Clinical Practice

The results of this study emphasize the need for clinicians to maintain a high index of suspicion for haematological malignancies in patients presenting with extreme leukocytosis, particularly when the WBC count exceeds 50×10^9^/L. Early identification and referral for appropriate diagnostic testing, including bone marrow biopsy and flow cytometry, are critical in improving patient outcomes. The presence of severe thrombocytopenia, splenomegaly, and lymphadenopathy should further raise suspicion of malignancy.

### Study Limitations

This study has several limitations. First, advanced diagnostic tests such as flow cytometry and molecular assays were not universally available, potentially leading to the under-diagnosis of certain haematological malignancies. Second, many patients in the infection group lacked microbiological culture data, limiting our ability to define the infectious etiologies precisely. Future studies should incorporate comprehensive diagnostic testing to provide a more accurate delineation of EL etiologies.

## Conclusion

In conclusion, this study demonstrates that extreme leukocytosis with WBC counts ≥50×10^9^/L is more commonly associated with malignant etiologies, particularly haematological malignancies, than infections. Clinicians should remain vigilant for malignancies in patients with EL and perform appropriate diagnostic testing to guide management. Early diagnosis and intervention are crucial to improving outcomes for patients with extreme leukocytosis.

## Data Availability

All data produced in the present study are available upon reasonable request to the authors.

## Notes

### Competing Interest Statement

The authors have declared no competing interest.

### Funding Statement

This study did not receive any funding

### Author Declarations

Mbeya Medical Research Ethics Committee of Mbeya Zonal Referral Hospital gave ethical approval for this work (Ref No: SZEC-2439/R.A/24/01)

